# Combined detrimental effect of male sex and GBA1 variants on cognitive decline in Parkinson’s Disease

**DOI:** 10.1101/2024.04.02.24305191

**Authors:** Silvia Paola Caminiti, Micol Avenali, Alice Galli, Rachele Malito, Giada Cuconato, Andrea Pilotto, Alessandro Padovani, Fabio Blandini, Daniela Perani, Cristina Tassorelli, Enza Maria Valente, Parkinson’s Progression Markers Initiative (PPMI)

## Abstract

**Background and Objective:** Heterozygous variants in the glucocerebrosidase gene (*GBA1*) are the major genetic risk factor for Parkinson’s Disease (PD). GBA-PD has been associated with worse progression and higher risk of cognitive decline. Here we took advantage of the Parkinson’s Progression Markers Initiative (PPMI) to investigate whether sex could interact with *GBA1* carrier status in determining the clinical phenotype, with a special focus on cognitive decline.

**Methods:** We evaluated 118 PD subjects carrying *GBA1* variants (GBA-PD) and 450 with wild-type alleles (nonGBA-PD) included in the PPMI. Dopaminergic activity was assessed in a subset of 248 subjects (65%) with available ^123^I-FP-CIT SPECT scans. Clinical features and dopaminergic activity were investigated in GBA-PD vs. nonGBA-PD groups, upon stratification by sex. PD subjects were followed for up to 6.5 years (median 6 years). Cox regression was used to model the hazard ratio (HR) of (1) *GBA1* genotype, (2) sex, (3) gene-by-sex interaction on cognitive decline at follow-up.

**Results:** Regardless of genotype, men suffering from PD exhibited higher motor disability while women showed more autonomic dysfunction. At baseline, GBA-PD showed more severe motor and non-motor features, and reduced dopamine uptake in the bilateral ventral putamen compared to nonGBA-PD. Within the GBA-PD group, males had higher occurrence of REM sleep behavior disorder and memory deficits. Of note, GBA-PD females showed a greater striatal dopaminergic deficit compared to males, despite presenting similar motor impairment. In longitudinal assessment, Cox Regression revealed that male sex (HR = 1.7), *GBA1* carrier status (HR =1.6) and, most importantly, GBA-by-male sex interaction (HR = 2.3) were significantly associated with a steeper cognitive decline. Upon stratification for *GBA1* variant class, both “severe” and “mild” variants were associated with increased risk of cognitive decline, again more relevant in males (HR = 2.3).

**Discussion:** We show, for the first time, that male sex and *GBA1* carrier status have an additive value in increasing the risk of cognitive decline in PD, despite the heightened dopaminergic vulnerability observed in GBA-PD females. The effect of sex on *GBA1*-related pathology warrants further examination and should be considered in future trials design and patients’ selection.

## 1. Introduction

Parkinson’s disease (PD) is a neurodegenerative condition characterized by a heterogeneous range of motor and non-motor symptoms^1^. Among these, the occurrence of cognitive impairment is one of the main causes of a poorer quality of life^2^. The different clinical trajectories observed in PD, especially as regards to non-motor symptoms, pose considerable challenges in identifying appropriate participants for clinical trials. As a result, there is a pressing need to a more accurate identification of factors that may increase the risk of PD subjects to develop cognitive impairment. Genetic factors are known to contribute to the susceptibility to cognitive decline and dementia in PD^3^. Heterozygous variants in the glucocerebrosidase gene (*GBA1*), occurring in about 8-12% of PD subjects worldwide, seem to accelerate the neurodegenerative process already in the earlier stages of the disease^4^, leading to a significant dopaminergic damage and a more severe clinical phenotype. Indeed, GBA-PD subjects have generally an earlier disease onset, a higher prevalence of non-motor symptoms and a greater risk of progression to dementia compared to non-mutated subjects^5^.

Sex is another established factor that affects incidence, natural history, and phenotype of the disease, as suggested by a recent meta-analysis showing a clear male preponderance over females (59% vs 41%)^6^. Furthermore, men manifest on average an earlier age at disease onset, more severe motor symptoms, and faster disease progression than women^7^. From a pathogenetic perspective, genetic, hormonal, neuroendocrinal and molecular players all contribute towards these sex-related differences^8^. In particular, steroid sex hormones, and especially oestrogens, seem to play a crucial neuroprotective role and anti-inflammatory function in PD^4^.

Notably, *GBA1* pathogenic variants are equally detected in PD subjects of both sexes, surpassing the potential impact of environmental exposures and hormonal influences that likely result in the higher male prevalence characterizing idiopathic PD^4^. In this context, it becomes crucial to investigate the potential interactions between *GBA1* genotype and sex, and their combined influence on clinical trajectories in PD.

In this study, we embarked in a comprehensive exploration of the mutual role of sex and *GBA1* mutations in modulating dopaminergic vulnerability as well as the risk to develop dementia in PD subjects. By shedding light on these multifaceted dimensions, we aim to deepen our understanding of PD cognitive manifestations and pave the way for more targeted and effective interventions.

## 2. Materials and Methods

### 2.1 Subjects

This analysis used data openly available from Parkinson’s Progression Markers Initiative (PPMI) database (www.ppmi-info.org/data), a multicentre, prospective, longitudinal study that aims to identify genetic, blood, cerebral spinal fluid, and imaging biomarkers of PD progression. Data used in the preparation of this article were obtained [between Jul 1, 2010, and Jun 1, 2019] from the Parkinson’s Progression Markers Initiative (PPMI) database (www.ppmi-info.org/access-data-specimens/download-data), RRID:SCR_006431. For up-to-date information on the study, visit www.ppmi-info.org.

The enrolment criteria for PD participants in the PPMI included the following conditions: age older than 30 years, a diagnosis of PD within 2 years prior to the screening visit, the presence of asymmetric resting tremor or asymmetric bradykinesia, or meeting two of the following criteria: bradykinesia, rigidity, and resting tremor. Additionally, participants were required to be untreated for PD at the baseline visit, to be in Hoehn and Yahr stage 1 or 2, and to exhibit a dopamine transporter deficit on imaging.

We included 568 PD subjects (mean age in years ± SD: 60.18 ± 10.1; sex [F/M]: 232/336) and with a clinical follow-up ≥12 months (mean ± SD: 77 ± 3.2 months; [min/max: 1/12 years]. Among them, 118 (20.8%) carried a GBA1 variant (GBA-PD), while the remaining 487 (79.2%) were GBA-negative (nonGBA-PD). Moreover, 133 subjects (23.4%) carried the ApoE D4 allele, of whom 10 (1.8%) in homozygosity. Of the 118 GBA-PD subjects, 23 (19.5%) had at least one ApoE D4 allele, of which 4 (3.4%) were ApoE D4/D4 homozygotes.

From this included sample, 248 subjects (GBA-PD/non-GBA-PD: 43/205) were acquired at baseline with ^123^I-FP-CIT-SPECT to image dopamine transporter (DAT) binding.

The institutional review board approved the study at each site, and the participants provided written informed consent.

### 2.2 Genotyping

DNA was extracted from whole blood according to the study protocol described in the PPMI Biological Products Manual (http://ppmi-info.org/study-design). Exons 1-11 of the *GBA1* gene were Sanger sequenced. Variants were subdivided in the following classes^11^: “risk” (E365K, T408M), “mild” (E365K/N409S, N409S, N409S/N409S, R535H), “severe” (E365K/N431S/L483P, E427K/L483P, IVS2+1G>A, L29Afs*18, L483P, R502C, T408M/R159W) and “unknown” (A495P, G154R/G232E, I528L, K13R, R78C, R83C, R83C/N409S).

ApoE genotypes (ε2, ε3 and ε4 alleles) were obtained by genotyping two single nucleotide polymorphisms (SNPs) (rs429358 and rs7412) by TaqMan assay on the NeuroX genotyping platform^12^. According to the presence/absence of the ApoE D4 allele, participants were divided into ApoE D4 Het (heterozygous for the D4 allele), ApoE ε4 Hom (homozygous for the D4 allele), and ApoE ε4 negative (carrying D2/D2, D2/D3 or D3/D3 genotypes).

### 2.3 Clinical assessment

We included the Movement Disorders Society-Unified Parkinson’s Disease Rating Scale-Part III (MDS-UPDRS-III)^13^ to assess motor function. Non-motor clinical assessments included the Rapid Eye Movement (REM) Sleep Behaviour Disorder Questionnaire (RBDSQ)^15^ to evaluate sleep behaviour, the Scale for Outcomes in PD-Autonomic (SCOPA-AUT)^16^ to explore autonomic dysfunction and the Geriatric Depression Scale (GDS) for the assessment of depressive symptoms. Global cognition was tested with the Montreal Cognitive Assessment (MoCA)^17^, adjusted for age and education. We obtained measures of MoCA deflection, namely the number of MoCA points lost per year, by considering both baseline and last follow-up MoCA score available. Moreover, subjects with a MoCA score ≤ 26 at last follow-up were classified as affected by cognitive impairment^18^.

We also collected neuropsychological measures evaluating cognitive domains usually considered particularly impaired in PD, including t-score of Hopkins Verbal Learning Test-Revised (HLVT-R)^19^ [for total recall, delayed recall, and recognition-discrimination] to assess memory; scores corrected for age and education of the Benton Judgment of Line Orientation (BJLO) 15-item version to assess visuospatial function ^20^; and the scaled scores of Letter-Number Sequencing (LNS)^21^ and t-score of semantic fluency^22^ to assess executive skills and working memory.

### 2.4 ^123^I-FP-CIT-SPECT acquisition and preprocessing

We retrieved reconstructed imaging data related to ^123^I-FP-CIT-SPECT from the PPMI website. Images were acquired using Siemens or General Electric SPECT tomographs, approximately 3-4 hours after administration of the ^123^I-FP-CIT tracer. The imaging protocol used for PPMI scans has been previously described^9,10^.

Preprocessing of SPECT brain images was conducted using Statistical Parametric Mapping software (SPM12, Wellcome Trust Centre for Neuroimaging, London, UK, available at: https://www.fil.ion.ucl.ac.uk/spm/), run with MATLAB R2022b (MathWorks Inc., Sherborn, MA, USA). First, each image was spatially normalized to a high-resolution ^18^F-DOPA template (http://www.nitrc.org/projects/spmtemplates)^10^ using the old normalize function in SPM12. Parametric binding potential were generated for each subject and voxel-wise using the Image Calculator (ImCalc) function in SPM12. The superior lateral occipital cortex was considered as the reference background region^23^.

Calculation of the asymmetry index (AI) was conducted following the standard formula, as described^23^.

### 2.5 Brain Dopaminergic Activity

A Voxel-wise multiple linear regression model was employed in SPM12 to compare GBA-PD and nonGBA-PD. The model included age at acquisition, AI and MDS-UPDRS-III as covariates. We set our voxel-wise significance threshold at p < 0.05 (uncorrected) and a minimum cluster extent of 100 voxels. Cluster-corrected statistical maps were saved as NifTI files. The resulting cluster obtained in the previous step was, then, transformed into a binary mask and used to extract parametric binding potentials from each group. We compared the distributions of binding potentials across the four clinical groups through non-parametric ANCOVA with post-hoc Bonferroni correction, including age at acquisition, AI and MDS-UPDRS-III as covariates.

### 2.6 Statistical Analysis

Demographic and clinical features were first compared between GBA-PD and nonGBA-PD, and then in subgroups stratified by sex, by means of ANOVA for continuous variables and Chi-squared tests for categorical variables. The effect of *GBA1* variants on cognitive deflection was tested via ANOVA. Differences in sex distribution in subjects stratified by *GBA1* variant type were determined using one-sample tests of equality of proportions.

Cox regression models were used to model the effect of (1) genetics, (2) sex, and (3) interaction between the considered variables on cognitive decline at follow-up. The Cox Regression Models were adjusted for disease duration and education. When we evaluated the effect of *GBA*1 and ApoE ε4 mutations on the risk of cognitive decline we added sex as confounding variable.

We tested a model including only confounders; if the output was statistically significant, the Cox proportional hazards models were adjusted for confounders by time-dependent on sex and genetics variables.

Hazard ratios (HRs) and 95% confidence intervals (CIs) were calculated, respectively. Significance threshold p < 0.05 was established for all tests.

All statistical analyses were performed by means of Statistical Package for Social Sciences (SPSS) version 28 and Python packages.

### 2.7 Standard Protocol Approvals, Registrations, and Patient Consents

Each PPMI participating site received approval from their local ethic committee before study initiation, and written informed consent was obtained from all participants before enrolment. We have obtained permission for publishing our research from the Data and Publication Committee of the PPMI study.

### 2.8 Data Availability

All the data used in this study are publicly available in the PPMI repository (www.ppmi-info.org/access-data-specimens/download-data).

## 3. Results

### 3.1 Baseline features

The demographic and clinical characteristics of the whole GBA-PD (M/F: 59/59) and nonGBA-PD (M/F: 277/173) groups are shown in **Table 1**. Upon *GBA1* variant classification, 23.7% subjects carried “risk” variants, 55.9% “mild”, 11.9% “severe”, and 8.5% “unknown”. The p.N409S (N370S) mild variant emerged as predominant, being present in 50% of mutated individuals (**Supplementary Table 1**).

**Table 1.**
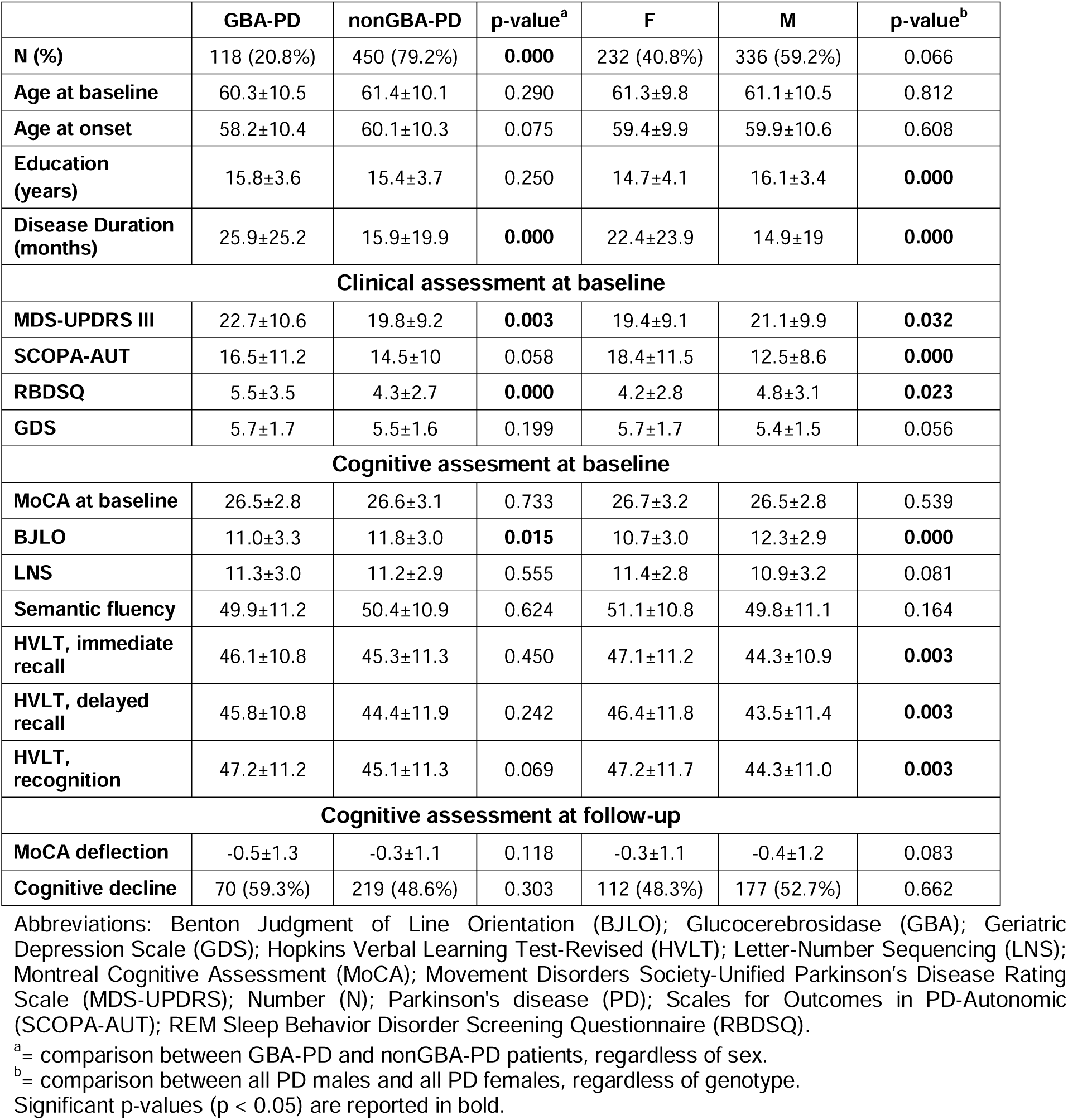
Demographic and clinical features of GBA-PD and nonGBA-PD groups.

At baseline, the two groups were comparable for age and educational level. GBA-PD showed longer disease duration, a significantly more severe motor profile and more prevalent REM sleep disorders than nonGBA-PD. On the cognitive side, MoCA scores were comparable in the two groups, although GBA-PD showed lower performance on visuospatial function (BJLO).

When assessing sex-related differences, regardless of *GBA1* carrier status, PD males showed more severe RBD and motor symptoms, while females showed greater autonomic dysfunction. PD males also showed lower memory performance than females, while the opposite was observed for visuospatial scores, with no differences in MoCA scores between sexes.

Demographic and clinical data upon stratification by sex and genotype are shown in **Table 2**. GBA-PD males had significantly more severe REM sleep disturbances than all other groups, and also the worst motor scores (albeit significant only vs nonGBA-PD females). Other inter-group comparisons at baseline confirmed the former observations of worse autonomic dysfunction, worse visuospatial performance and better memory performance in females compared to males.

**Table 2.** Demographic and clinical features of GBA-PD and nonGBA-PD groups divided by sex.

|  | **GBA-PD**<br>**F** | **GBA-PD**<br>**M** | **nonGBA-PD**<br>**F** | **nonGBA-PD**<br>**M** | **p-**<br>**value** | **Post hoc** |
| --- | --- | --- | --- | --- | --- | --- |
| **N (%)** | 59 (10.4%) | 59 (10.4%) | 173 (30.4%) | 277 (48.8%) | **0.000** |  |
| **Age at baseline** | 60.8±10.3 | 59.7±10.5 | 61.5±9.6 | 61.4±10.4 | 0.690 |  |
| **Age at onset** | 58.0±10.6 | 58.0±11.2 | 59.7±9.8 | 60.1±10.5 | 0.299 |  |
| **Education**<br>**(years)** | 15.3±4.1 | 16.3±2.9 | 14.4±4 | 16.0±3.5 | **0.000** | nonGBA-PD F < GBA-PD M<br>nonGBA-PD F < nonGBA-PD M |
| **Disease Duration**<br>**(months)** | 28.1±24.2 | 23.7±25.2 | 20.5±23.2 | 13.1±16.9 | **0.000** | nonGBA-PD M < GBA-PD F<br>nonGBA-PD M < GBA-PD M<br>nonGBA-PD M < nonGBA-PD F |
| **Clinical assessment at baseline** |  |  |  |  |  |  |
| **MDS-UPDRS III** | 21.5±9.6 | 24.0±11.4 | 18.6±8.8 | 20.5±9.4 | **0.002** | GBA-PD M > nonGBA-PD F |
| **SCOPA-AUT** | 18.8±11.1 | 14.2±10.7 | 18.3±11.5 | 12.1±8.0 | **0.000** | GBA-PD F > nonGBA-PD M<br>nonGBA-PD F > GBA-PD M<br>nonGBA-PD F > nonGBA-PD M |
| **RBDSQ** | 4.5±3 | 6.5±3.7 | 4.1±2.7 | 4.4±2.8 | **0.000** | GBA-PD M > GBA-PD F<br>GBA-PD M > nonGBA-PD F<br>GBA-PD M > nonGBA-PD M |
| **GDS** | 5.9±1.9 | 5.5±1.4 | 5.6±1.7 | 5.4±1.58 | 0.146 | - |
| **Cognitive assesment at baseline** |  |  |  |  |  |  |
| **MoCA (at**<br>**baseline)** | 26.9±2.9 | 26.2±2.6 | 26.6±3.8 | 26.6±2.8 | 0.638 | - |
| **BJLO** | 10.2±3.1 | 10.9±2.2 | 10.9±3.0 | 12.4±2.9 | **0.000** | GBA-PD F < GBA-PD M<br>GBA-PD F < nonGBA-PD M<br>nonGBA-PD F < GBA-PD M<br>nonGBA-PD F < nonGBA-PD M |
| **LNS** | 11.3±3.0 | 11.4±3.0 | 10.8±3.2 | 11.4±2.8 | 0.234 | - |
| **Semantic**<br>**Fluency** | 49.5±11.5 | 50.2±11.0 | 51.6±10.5 | 49.7±11.0 | 0.286 | - |
| **HVLT, immediate**<br>**recall** | 47.9±11.3 | 44.4±10.1 | 46.8±11.1 | 44.3±11.2 | **0.026** | - |
| **HVLT, delayed**<br>**recall** | 48.6±10.4 | 43.0±10.4 | 45.7±12.2 | 43.6±11.6 | **0.009** | GBA-PD M < GBA-PD F<br>nonGBA-PD M < GBA-PD F |
| **HVLT,**<br>**recognition** | 50.0±10.1 | 44.4±11.6 | 46.3±12.1 | 44.3±10.9 | **0.003** | GBA-PD M < GBA-PD F<br>nonGBA-PD M < GBA-PD F |
| **Cognitive assessment at follow-up** |  |  |  |  |  |  |
| **MoCA deflection** | -0.1±0.8 | -0.9±1.5 | -0.3±1.1 | -0.3±1.1 | **0.001** | GBA-PD M < GBA-PD F<br>GBA-PD M < nonGBA-PD F<br>GBA-PD M < nonGBA-PD M |
| **Cognitive decline** | 30 (50.8%) | 40 (66.8%) | 82 (47.4%) | 137 (49.5%) | 0.219 |  |

Abbreviations: Beck Depression Inventory (BDI); Benton Judgment of Line Orientation (BJLO); Glucocerebrosidase (GBA); Hopkins Verbal Learning Test-Revised (HVLT); Letter-Number Sequencing (LNS); Montreal Cognitive Assessment (MoCA); Movement Disorders Society-Unified Parkinson's Disease Rating Scale (MDS-UPDRS); Number (N); Parkinson's disease (PD); Scales for Outcomes in PD-Autonomic (SCOPA-AUT); REM Sleep Behavior Disorder Screening Questionnaire (RBDSQ).
\* (Post Hoc) = Significance did not withstand Bonferroni's correction. Significant p-values (p < 0.05) are reported in bold.

There was no difference in sex-specific distribution of *GBA1* carriers (59 men, 50%) and of variants subgroups (“Risk” = 13 men, 46.4%; “Mild” 34 men, 51.5%; “Severe” 6 men, 42.8%) **(Supplementary Figure 1)**, but selected variants prevailed in either sex (e.g. T408M in males and E365K in females). The different subgroups defined by the genetic variants were again similar in terms of MoCA scores at baseline. Heterozygous and homozygous ApoE ε4 genotypes were significantly more frequent in GBA-PD males than females (**Supplementary Table 1**).

### 3.2 Dopamine Transporter Imaging at baseline

In accordance with our previous report^23,24^, GBA-PD showed reduced dopamine uptake in bilateral ventral putamen (p<0.05 at cluster level and p=0.01 at peak level) as compared to nonGBA-PD. Despite comparable motor clinical profiles among subgroups (**Table 3**), GBA-PD females showed the greatest dopaminergic impairment, which was significantly worse than both males and females nonGBA-PD subjects (**Figure 1**).

**Figure 1.**
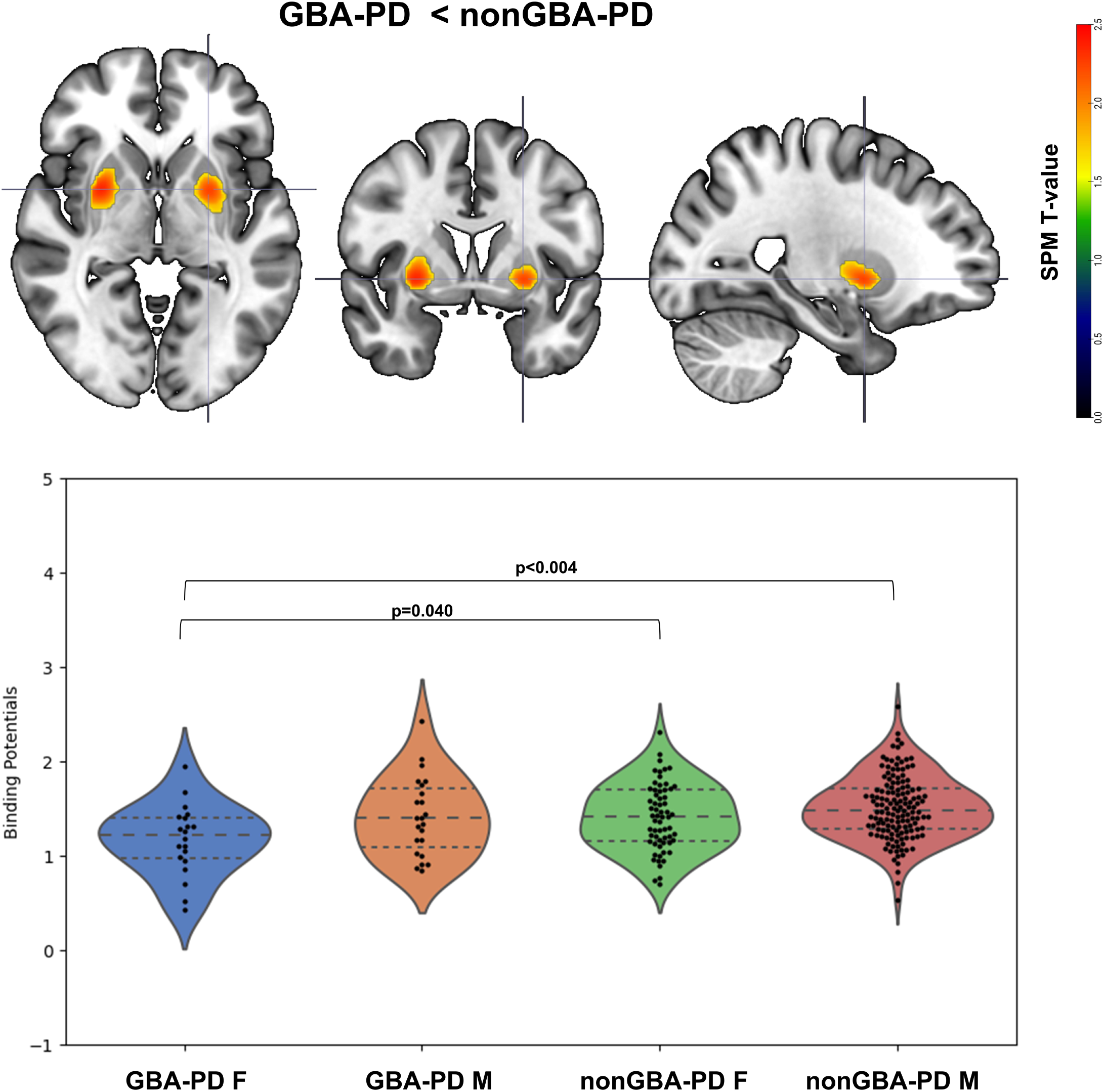
Dopaminergic uptake differences between groups. (Top) Significantly reduced dopamine binding in GBA-PD subjects compared to nonGBA-PD subjects, resulting from the voxel-wise regression model. (Bottom) Violin plots representing significant differences in dopaminergic binding potentials in bilateral ventral putamen among the four clinical groups.

**Table 3.** Demographic and clinical features of GBA-PD and nonGBA-PD subgroups with ^123^I-FP-CIT-SPECT data available, divided by sex.

|  | GBA-PD<br>F | GBA-PD<br>M | nonGBA-PD<br>F | nonGBA-PD<br>M | p-value | Post hoc |
| --- | --- | --- | --- | --- | --- | --- |
| **N (%)** | 20 (33.9%) | 23 (39%) | 63 (36.4%) | 142 (51.3%) | 0.217 |  |
| **Age at SPECT** | 63.6±9.7 | 63.7±11.2 | 61.5±7.3 | 64.7±7.4 | 0.277 |  |
| **Education**<br>**(years)** | 15.4±5.1 | 16.7±3.0 | 15.2±2.7 | 15.6±3.0 | 0.263 | - |
| **Disease Duration**<br>**(months)** | 33.5±26.8 | 27.7±28.1 | 8.0±8.7 | 6.6±6.3 | **0.000** | nonGBA-PD M < GBA-PD F<br>nonGBA-PD M < GBA-PD M<br>nonGBA-PD M < nonGBA-PD F<br>nonGBA-PD F < GBA-PD F |
| **AI** | 5.0±3.4 | 4.9±3.4 | 7.1±3.9 | 5.2±3.5 | **0.003** | nonGBA-PD M < nonGBA-PD F |
| **Clinical assesment at baseline** |  |  |  |  |  |  |
| **MDS-UPDRS III** | 20.9±10.7 | 23.8±11.4 | 20.3±8.4 | 21.4±8.9 | 0.469 | - |
| **SCOPA-AUT** | 17.3±10.5 | 13.9±8.2 | 17.3±10.1 | 12.8±8.0 | **0.004** | nonGBA-PD F > nonGBA-PD M |
| **RBDSQ** | 3.5±2.1 | 7.2±3.9 | 3.8±2.6 | 4.7±3.0 | **0.000** | GBA-PD M > GBA-PD F<br>GBA-PD M > nonGBA-PD F<br>GBA-PD M > nonGBA-PD M |
| **GDS** | 5.9±1.7 | 5.6±1.4 | 5.3±1.3 | 5.2±1.5 | 0.115 | - |
| **Cognitive assesment at baseline** |  |  |  |  |  |  |
| **MoCA (at**<br>**baseline)** | 27.4±2.2 | 26.0±2.6 | 27.5±1.9 | 26.6±2.6 | **0.016** | GBA-PD M < nonGBA-PD F |
| **BJLO** | 10.6±2.8 | 12.9±2.2 | 11.8±2.5 | 12.4±2.8 | **0.016** | GBA-PD F < GBA-PD M<br>GBA-PD F < nonGBA-PD M |
| **LNS** | 11.6±3.2 | 11.3±2.9 | 11.8±2.8 | 11.4±2.7 | 0.825 | - |
| **Semantic**<br>**Fluency** | 50.4±10.7 | 53.2±8.3 | 51.2±9.7 | 50.6±9.0 | 0.646 | - |
| **HVLT, immediate**<br>**recall** | 50.8±12.4 | 46.7±8.8 | 49.4±9.5 | 42.2±11.1 | **0.000** | nonGBA-PD M < GBA-PD F<br>nonGBA-PD M < nonGBA-PD F |
| **HVLT, delayed**<br>**recall** | 49.4±11.6 | 43.7±11.0 | 47.6±10.5 | 42.0±11.9 | **0.002** | nonGBA-PD M < GBA-PD F<br>nonGBA-PD M < nonGBA-PD F |
| **HVLT,**<br>**recognition** | 52.4±7.3 | 46.0±8.2 | 47.1±11.6 | 42.8±11.2 | **0.001** | nonGBA-PD M < GBA-PD F |
| **Cognitive assessment at follow-up** |  |  |  |  |  |  |
| **MoCA deflection** | -0.2±1.1 | -0.7±1.4 | -0.4±1.3 | -0.4±0.9 | 0.490 | - |
| **Cognitive decline** | 9 (45%) | 15 (65.2%) | 27 (43%) | 86 (60.6%) | 0.075 |  |

Abbreviations: Asymmetry Index (AI); Beck Depression Inventory (BDI); Benton Judgment of Line Orientation (BJLO); Glucocerebrosidase (GBA); Hopkins Verbal Learning Test-Revised (HVLT); Letter-Number Sequencing (LNS); Montreal Cognitive Assessment (MoCA); Movement Disorders Society-Unified Parkinson's Disease Rating Scale (MDS-UPDRS); Number (N); Parkinson's disease (PD); Scales for Outcomes in PD-Autonomic (SCOPA-AUT); REM Sleep Behavior Disorder Screening Questionnaire (RBDSQ).
^ (Post Hoc) = Significance did not withstand Bonferroni's correction. Significant p-values (p < 0.05) are reported in bold.

### 3.3 Cognitive trajectories at follow-up

During the follow-up period (6.5 ± 3.2 years), 289 out of 568 PD patients (50.9%) experienced cognitive deterioration, with MoCA scores ≤ 26 at follow-up assessment; this subgroup included 70 GBA-PD patients (out of 118 = 59.3%). *GBA1* carrier status produced a significant risk of cognitive decline over time (HR = 1.594, 95% CI = 1.208 – 2.105, p = 0.001), which did not change when the model was adjusted for sex (HR = 1.705, 95% CI = 1.288 – 2.258, p < 0.001). Consistent with these predictions, the levels of MoCA deflection were significantly higher in GBA-PD (-0.51 point/year) compared to nonGBA-PD (-0.33 point/year).

Among GBA-PD subjects manifesting cognitive decline at follow-up, 12 (17.1%, 9/3 M/F) showed at least one ApoE □4 allele, and 2 (2.8%, 2 M) had a homozygous ApoE □4 genotype (**Supplementary Table 1**). While the interaction between *GBA1* carrier status and the presence of at least one ApoE □4 allele did not reach statistical significance, the lack of both *GBA1* and APoE □4 mutations resulted in a protective effect on cognitive decline (HR = 0.791, 95% Cl = 0.624 – 1.003, p = 0.05).

Both “severe” and “mild” *GBA1* variants were significantly associated with conversion to cognitive decline, with a frequency of 71.4% and 62.1%, respectively, while subjects carrying “risk” (53.7%) and “unknown” (40%) variants had lower percentage of subjects manifesting cognitive decline (**Supplementary Table 1**). Consistently, Cox regression showed that GBA-PD subjects carrying either “severe” or “mild” variants had an increased risk of cognitive decline (HR = 1.865, 95% Cl = 1.351 – 2.575, p < 0.001), while “risk” and “unknown” variants showed no significant effect on the PD cognitive trajectory.

Notably, when assessing the combined effect of *GBA1* genotype and sex, GBA-PD males showed a significantly steeper MoCA deflection (-0.90 point/year) not only when compared to GBA-PD females (-0.13 point/year), but also when compared to nonGBA-PD subjects, both males (-0.34 point/year) and females (-0.31 point/year) (**Supplementary Figure 2**). Accordingly, the interaction between male sex and *GBA1* carrier status significantly impacted on the risk of cognitive decline (HR=2.286, 95% CI = 1.623 - 3.222, p < 0.001).

When further stratifying for variant categories, an effect of sex on the risk of cognitive decline was found in carriers of both “severe” and “mild” variants, again with males showing significant higher risk of conversion than females (HR = 2.300, CI 95% = 1.218 – 4.341, p=0.010). All results are summarized in Survival Plots depicted in **Figure 2**.

**Figure 2.**
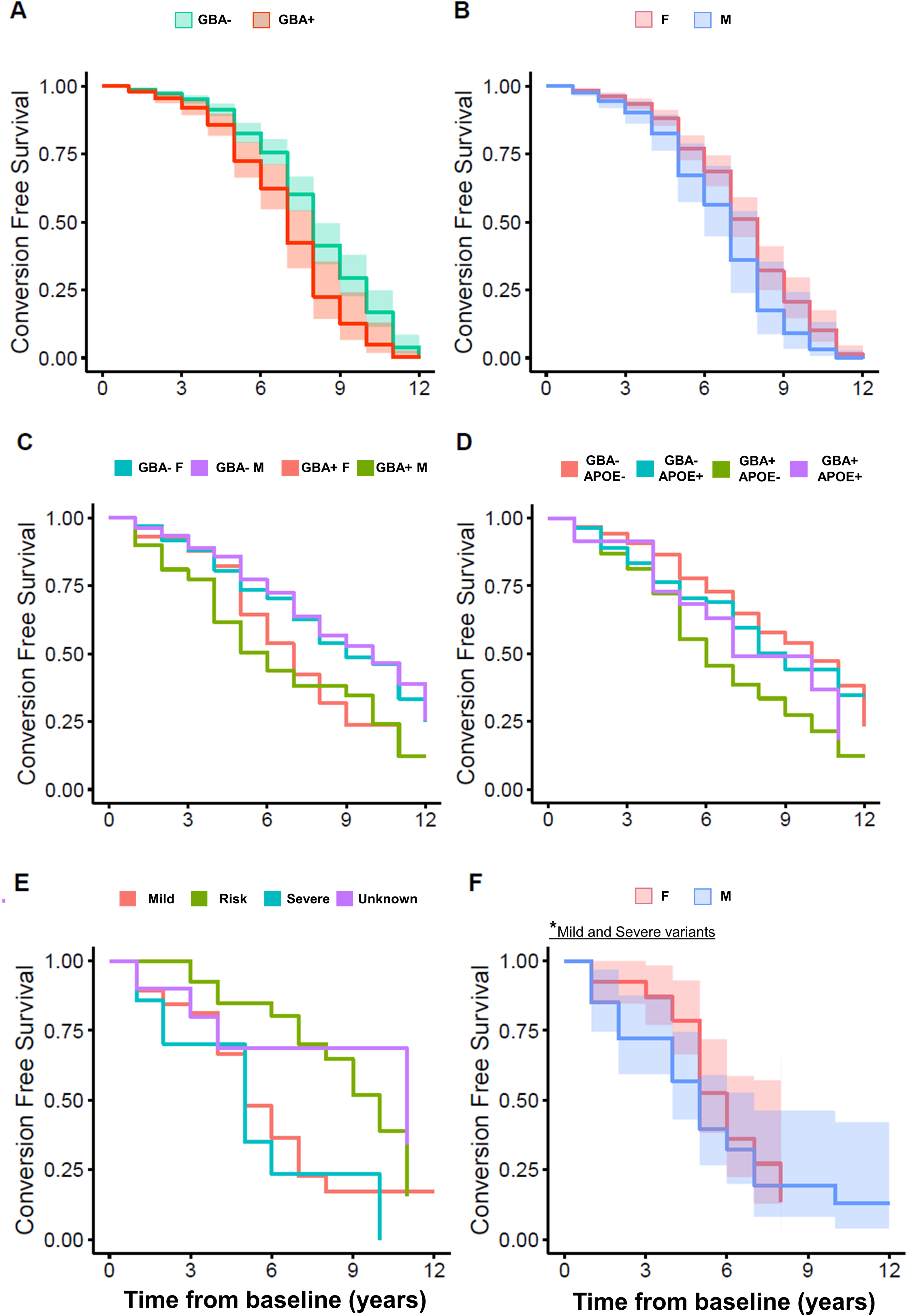
Survival plots for PD patients. Plots showing the association between the probability of conversion to cognitive deficits and the effect of (A) *GBA1* carrier status, (B) biological sex, (C) interaction between *GBA1* genotype and biological sex, (D) interaction between *GBA1* and ApoE ε4 carrier status, (E) GBA variants, (F) sex in carriers of mild and severe *GBA1* variants.

## 4. Discussion

The interaction between sex and genetics is complex and poorly understood in the context of PD^7^. Sex-related frequency differences have been reported in genetic forms of PD, with observed variation depending on the specific gene^25^. As regards *GBA1*, the sex distribution remains controversial^26–28^, with discrepancies depending on the *GBA1* variants under investigation^4,25^. For instance, a previous report indicated a preponderance of women among carriers of “severe” variants, while men were more likely to harbour “mild” and “risk” variants^25^. Here, in a large cohort of PD subjects extracted from the PPMI dataset, we failed to detect relevant differences in the prevalence of *GBA1* heterozygous carriers between men and women. Moreover, a comparable sex distribution was observed across the majority of *GBA1* variants, suggesting that observed differences are merely related to the different distribution of common *GBA1* variants across different populations and ethnicities.

The main aim of this study was to investigate the combined role of sex and *GBA1* carrier status in the risk of progression toward a cognitive decline, to address the key question whether *GBA1* mutations and sex have an independent or cumulative effect on cognitive outcomes. Our results highlighted, for the first time, that the combination of *GBA1* mutations and male sex is associated with a higher risk of cognitive impairment and a steeper cognitive decline along the disease course. This novel finding is in keeping with previous research reporting male sex as predictor of higher risk of developing cognitive decline^29^ or even dementia^30^, as well as with the known association of *GBA1* variants with cognitive impairment^24,31–33^. To the best of our knowledge, only a previous study reported similar evidence in a large cohort of 4,923 subjects with primary degenerative parkinsonism, finding that *GBA1* variants and male sex were associated with a higher proportion of subjects with PD-dementia and dementia with Lewy bodies than idiopathic PD^4^.

On the other hand, female biological sex seems to exert a protective effect also on GBA-PD condition. Indeed, we found no association between female sex and risk of cognitive decline in GBA-PD subjects, also supported by a slower cognitive decline in GBA-PD females than males. Taken together, these findings suggest the existence of relevant sex-related discrepancies in the manifestation of cognitive dysfunction in GBA-PD. Interestingly, despite a more benign clinical phenotype, GBA-PD females showed greater dopaminergic deficit as compared to GBA-PD males, suggesting that, in the course of the disease, GBA-PD females can counteract pathological brain changes through mechanisms of neural reserve and neural compensation^34^. In the general population, women tend to exhibit higher physiological levels of dopamine in the striatum, reflecting differences in basal dopamine system organization and/or neuroanatomy^35^. The dopamine system contains a high density of oestrogen receptors, through which hormones exert their protective role on dopaminergic functions^35^. Such protective effects of oestrogens are achieved by reducing oxidative stress and mitochondrial dysfunction, limiting neuroinflammation, and preventing the deposition of α-synuclein and neural injury^36^. Another aspect under investigation is related to the detrimental role of *GBA1* mutations on sphingolipid homeostasis, the latter found to be modulated by means of oestrogen receptor^37^. Thus, both environmental and hormonal factors may counteract PD-related pathology over the lifetime of pre-menopausal women, contributing to build a neural reserve through relevant neurobiological effects, even in *GBA1* carriers^38^. Overall, it is tempting to speculate that a more advanced stage of neurodegeneration is needed in females to reach the same clinical severity observed in GBA-PD males. Future prospective studies - focusing on the influence of hormones on *GBA1*-related pathology - could lead to a better understanding of the wide motor and cognitive between-sex variability in PD, as well as reveal new therapeutic avenues or preventive strategies.

Besides cognitive impairment, GBA-PD males showed higher occurrence of RBD disorders compared to all other groups. This finding is of particular interest as it is in line with the male predominance of RBD^39^, but also with the strong association between the presence of RBD and *GBA1* carrier status^24,40^. RBD in PD subjects is considered a marker of a more malignant phenotype, with more rapid progression of motor and non-motor symptoms^41^, as well as being related to more severe spreading of α-synuclein pathology at post-mortem assessment^42^.

The augmented risk of cognitive impairment may be also driven by the co-occurrence of *GBA1* mutations with the ApoE ε4 allele, which is by itself a strong risk factor for cognitive deterioration in Alzheimer’s disease^43^. A single study has so far demonstrated that carriers of both *GBA1* variants and ApoE ε4 alleles were at increased risk of a more severe course of PD^32^. Here, we do not find any significant risk, possibly due to the low statistical power – only 23 out of 118 PD bearing both mutations. Consistently, we found that the absence of both genetic mutations is protective against the development of cognitive decline in PD.

Another aspect emerging from our analysis is related to the differential effects of *GBA1* classes of variants on cognitive decline. Previous evidence showed conflicting results on the relationship between classes of *GBA1* mutations and PD phenotype as well as the risk for dementia^4,33,44,45^. Indeed, Cilia et al. (2016) found that carriers of “severe” mutations had greater risk for dementia compared to “mild” mutations, even if the latter showed a 2-fold higher risk of dementia than nonGBA-PD subjects^44^. Our former study on a large Italian cohort also showed that patients with “severe” GBA-PD exhibited greater risk of non-motor symptoms (e.g. cognitive impairment) compared to “mild” mutations^11^. Later on, Lunde et al. (2018) reported that “severe” variants are associated with a faster progression to dementia than carriers of “risk” polymorphisms (e.g. E365K)^33^, a finding not confirmed by Straniero et al. (2020), who found a higher risk of dementia not only in association with “severe” mutations but also with the E326K “risk” variant^4^.

The present study on the PPMI cohort confirms that *GBA1* “severe” variants are generally correlated with a more severe clinical phenotype, but it shows that “mild” variants also increase the risk of cognitive decline in PD. Conversely, a much lower proportion of “risk” variant carriers eventually converted to dementia at follow-up, suggesting that this category is characterized by a more benign outcome, especially on the cognitive side. The higher risk of conversion found in carriers of both “mild” and “severe” mutations may partially explain the considerable clinical variability reported among GBA-PD subjects in terms of cognitive dysfunction and motor disability^44^. However, we found that 38% of “mild” and 29% of “severe” variant carriers showed stable cognitive profiles at follow-up, suggesting that other risk factors, in combination with genetic risk factors, should be considered to shed light on PD heterogeneity. Overall, this still controversial evidence suggests that the current classification of *GBA1* variants, which is based on their role in Gaucher’s disease, may not adequately reflect their pathogenic role in PD, and new classification approaches should be investigated. Given these premises, future studies further addressing the issue of heterogeneity within the spectrum of *GBA1* genotypes and its relationship with sex should be implemented, also considering the effect on PD motor and non-motor phenotype and clinical trajectories. Moreover, our data should be confirmed in large population-based studies to limit bias in the ascertainment.

In conclusion, our results provide supporting evidence of the interplay between *GBA1* carrier status and sex in the progression of cognitive decline in a PD population. We confirm that *GBA1* variants are the major risk factor associated with cognitive impairment, however, this effect is particularly evident in association with the male sex. Indeed, we found that, among *GBA*1 carriers (mainly of “severe” and “mild” variants), PD males showed the greatest risk of develop cognitive impairment over time. These elements should be considered when interpreting the current literature and planning future studies. Understanding the role of genetic variants on the course of cognitive decline over PD progression will foster a more accurate disease prognosis and may help to a better future clinical trials design and patients’ selection. In particular, the effect of sex on *GBA1* mutation should be considered in the emerging therapeutic strategies targeting *GBA1*-regulated pathways^46^.

## Data Availability

All data produced in the present study are available upon reasonable request to the authors.

## Acknowledgment

Funding: PPMI – a public-private partnership – is funded by the Michael J. Fox Foundation for Parkinson’s Research and funding partners, including 4D Pharma, Abbvie, AcureX, Allergan, Amathus Therapeutics, Aligning Science Across Parkinson’s, AskBio, Avid Radiopharmaceuticals, BIAL, Biogen, Biohaven, BioLegend, BlueRock Therapeutics, Bristol-Myers Squibb, Calico Labs, Celgene, Cerevel Therapeutics, Coave Therapeutics, DaCapo Brainscience, Denali, Edmond J. Safra Foundation, Eli Lilly, Gain Therapeutics, GE HealthCare, Genentech, GSK, Golub Capital, Handl Therapeutics, Insitro, Janssen Neuroscience, Lundbeck, Merck, Meso Scale Discovery, Mission Therapeutics, Neurocrine Biosciences, Pfizer, Piramal, Prevail Therapeutics, Roche, Sanofi, Servier, Sun Pharma Advanced Research Company, Takeda, Teva, UCB, Vanqua Bio, Verily, Voyager Therapeutics, the Weston Family Foundation and Yumanity Therapeutics.

## Disclosure

The authors report no disclosures relevant to the manuscript.

Outside the submitted work, SPC is supported by #NEXTGENERATIONEU (NGEU) and funded by the Ministry of University and Research (MUR), National Recovery and Resilience Plan (NRRP), project MNESYS (PE0000006) – A multiscale integrated approach to the study of the nervous system in health and disease (DN. 1553 11.10.2022).

CT has received personal fees for participating in advisory boards for Eli Lilly.

| Appendix Authors |  |  |
| --- | --- | --- |
| Name | Location | Contribution |
| Silvia Paola Caminiti | Department of Brain and Behavioral Sciences, University of Pavia, Pavia, Italy | Design and conceptualization of the study; acquisition, analysis, and interpretation of the data; drafting of the manuscript |
| Micol Avenali | Parkinson and Movement Disorders Unit, IRCCS Mondino Foundation, Pavia, Italy | Interpretation of the data; drafting of the manuscript |
| Alice Galli | Neurology Unit, Department of Clinical and Experimental Sciences, University of Brescia, Italy | Acquisition, analysis, and interpretation of the data; drafting of the manuscript |
| Rachele Malito | Vita-Salute San Raffaele University, Milan, Italy | Acquisition, analysis, and interpretation of the data; drafting of the manuscript |
| Giada Cuconato | Department of Molecular Medicine, University of Pavia, Pavia, Italy | Acquisition, analysis, and interpretation of the data; critical revision |
| Andrea Pilotto | Neurology Unit, Department of Clinical and Experimental Sciences, University of Brescia, Italy | Interpretation of the data; critical revision of the manuscript |
| Alessandro Padovani | Neurology Unit, Department of Clinical and Experimental Sciences, University of Brescia, Italy | Interpretation of the data; critical revision of the manuscript |
| Fabio Blandini | IRCCS Ca' Granda Foundation, Ospedale Maggiore Policlinico, Milan, Italy | Interpretation of the data; critical revision of the manuscript |
| Daniela Perani | IRCCS San Raffaele Scientific Institute, Milan, Italy; Vita-Salute San Raffaele University, Milan, Italy | Interpretation of the data; critical revision of the manuscript |
| Cristina Tassorelli | Department of Brain and Behavioral Sciences, University of Pavia, Pavia, Italy | Interpretation of the data; critical revision of the manuscript |
| Enza Maria Valente | Neurogenetics Research Unit,<br>IRCCS Mondino Foundation, Pavia,<br>Italy | Design and conceptualization of the<br>study; interpretation of the data;<br>drafting of the manuscript |

