## Supplementary Figure 1 for "Combined detrimental effect of male sex and GBA1 variants on cognitive decline in Parkinson’s Disease"

**Supplementary Figure 1 - Sex distribution in the GBA-PD cohort**. Histogram depicting the percentage of males and females within the GBA-PD cohort. The y-axis indicates the numerosity of the specific subgroup.

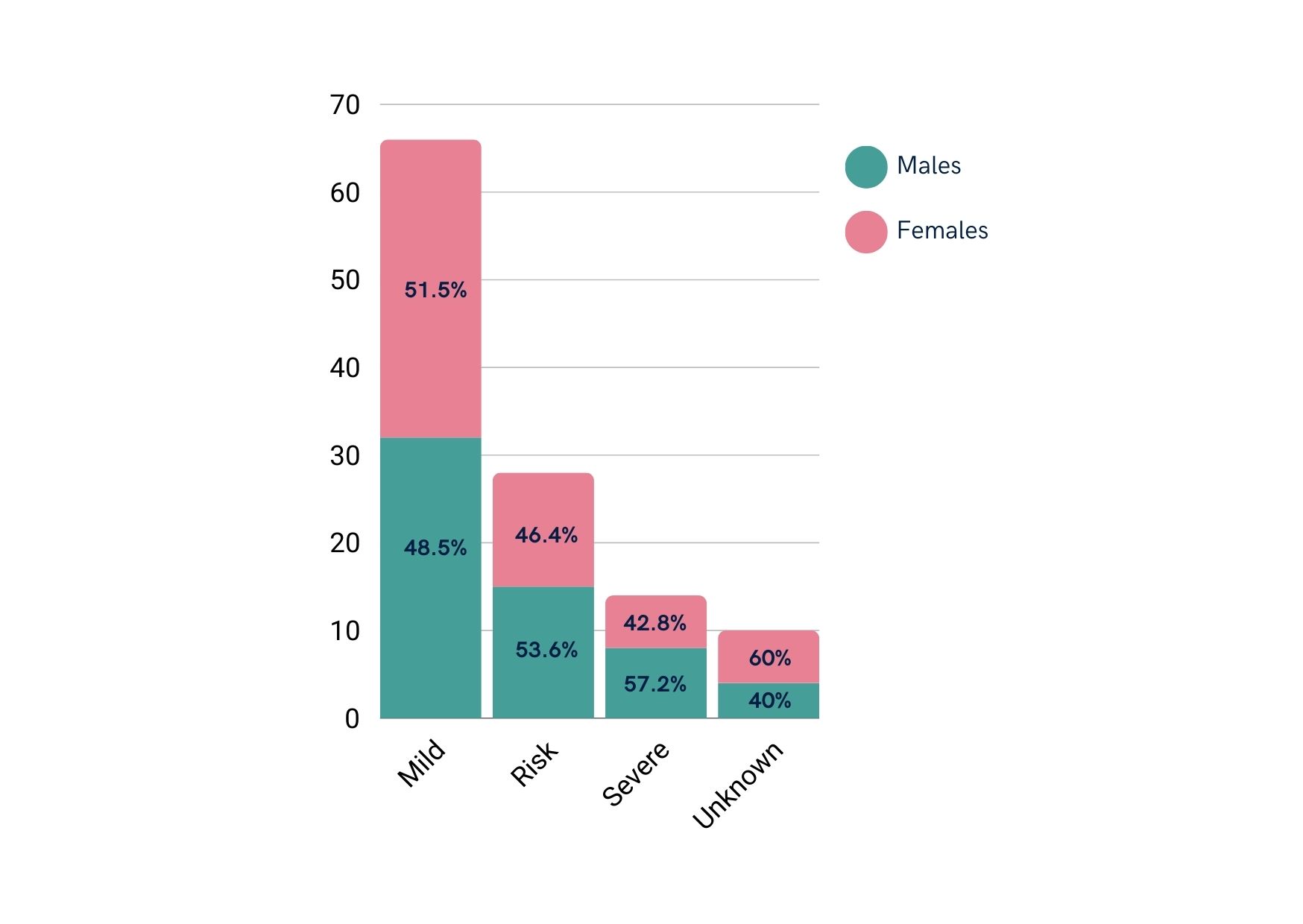
