## Supplementary Figure 2 for "Combined detrimental effect of male sex and GBA1 variants on cognitive decline in Parkinson’s Disease"

**Supplementary Figure 2** Cognitive trajectories (Montreal Cognitive Assessment total score) across the follow-up time in months in the four considered clinical groups. Thick lines indicate overall trend.

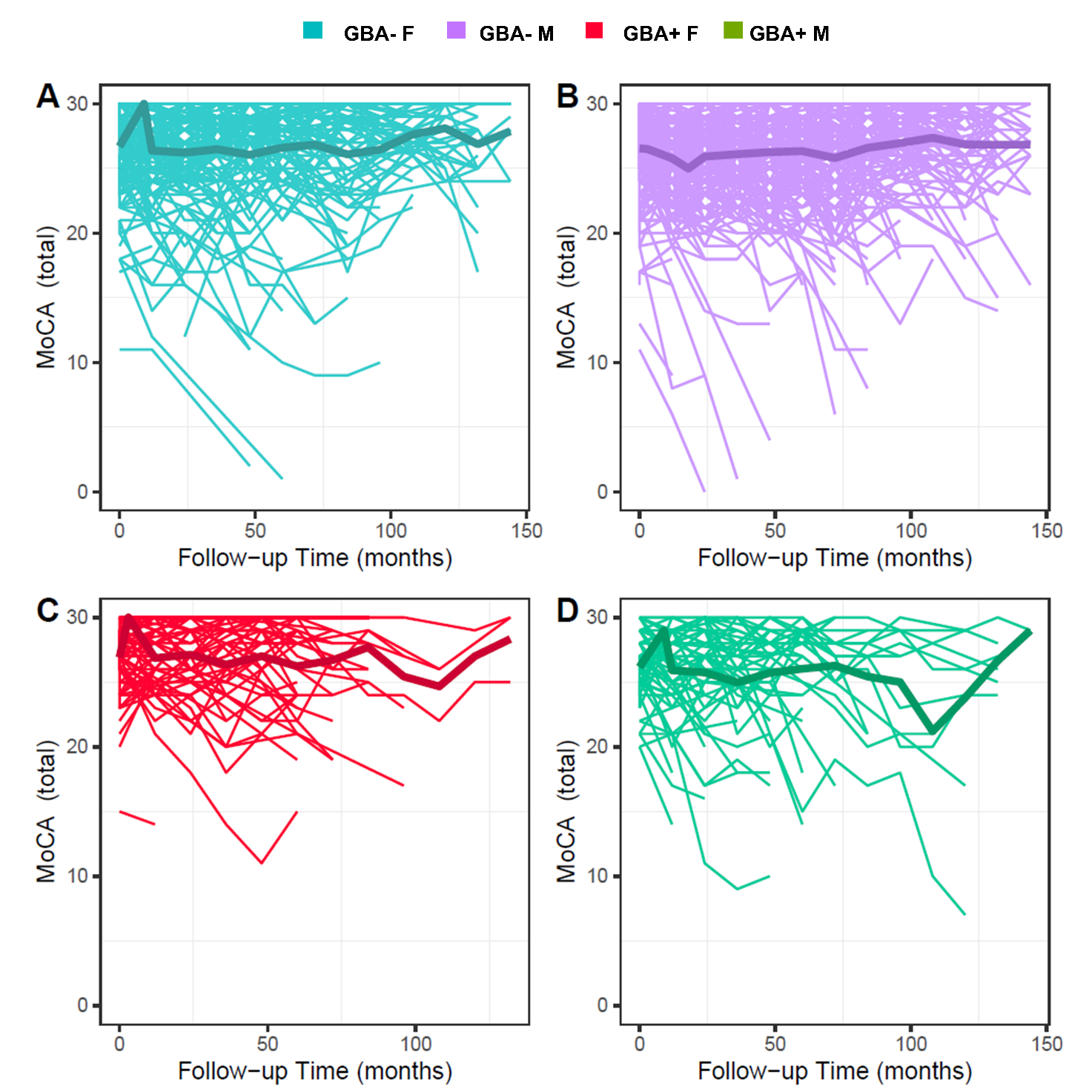
