## Supplementary Table 1 for "Combined detrimental effect of male sex and GBA1 variants on cognitive decline in Parkinson’s Disease"

**Supplementary Table 1** Distributions of genetic variants within the entire cohort of GBA-PD, as well as within subgroups characterized by cognitive impairment (CI) at follow-up, are presented, with stratification based on sex.

|  | **GBA-PD** | **GBA-PD F** | **GBA-PD M** | **GBA-PD**  **(CI)** | **GBA-PD F**  **(CI)** | **GBA-PD M**  **(CI)** | **p-value^a^** | **p-value^b^** |
| --- | --- | --- | --- | --- | --- | --- | --- | --- |
| **N** | 118 | 59 (50%) | 59 (50%) | 70 (59.3%) | 30 (42.8%) | 40 (57.1%) | 1.00 | 0.15 |
| **Risk** | 28 | 13 (46.4%) | 15 (53.6%) | 15 (53.7%) | 6 (40%) | 9 (60%) | 0.47 | **<0.05** |
| *E365K* | 18 | 11 (61.2%) | 7 (38.8%) | 9 (50%) | 5 (55.6%) | 4 (44.4%) | **0.02** | 0.26 |
| *T408M* | 10 | 2 (20%) | 8 (80%) | 6 (60%) | 1 (16.7%) | 5 (83.3%) | **< 0.001** | **< 0.001** |
| **Mild** | 66 | 34 (51.5%) | 32 (48.5%) | 41 (62.1%) | 19 (46.3%) | 22 (53.7%) | 0.76 | 0.46 |
| *E365K / N409S* | 1 | 1 (100%) | 0 (0%) | 0 (0%) | 0 (0%) | 0 (0%) | **< 0.001** | - |
| *N409S* | 59 | 31 (52.5%) | 28 (47.5%) | 38 (64.4%) | 18 (47.4%) | 20 (52.6%) | 0.61 | 0.6 |
| *N409S/N409S* | 5 | 1 (20%) | 4 (80%) | 3 (60%) | 1 (33.3%) | 2 (66.7%) | **< 0.001** | **< 0.001** |
| *R535H* | 1 | 1 (100%) | 0 (0%) | 0 (0%) | 0 (0%) | 0 (0%) | **< 0.001** | - |
| **Severe** | 14 | 6 (42.8%) | 8 (57.2%) | 10 (71.4%) | 4 (40%) | 6 (60%) | 0.15 | **<0.05** |
| *E365K / N431S / L483P* | 1 | 0 (0%) | 1 (100%) | 1 (100%) | 0 (0%) | 1 (100%) | **< 0.001** | **< 0.001** |
| *E427K / L483P* | 1 | 1 (100%) | 0 (0%) | 0 (0%) | 0 (0%) | 0 (0%) | **< 0.001** | - |
| *IVS2+1G>A* | 1 | 0 (0%) | 1 (100%) | 0 (0%) | 0 (0%) | 0 (0%) | **< 0.001** | - |
| *L29Afs*18* | 4 | 2 (50%) | 2 (50%) | 2 (50%) | 0 (0%) | 2 (100%) | 1.00 | **< 0.001** |
| *L483P* | 6 | 3 (50%) | 3 (50%) | 5 (83.3%) | 3 (60%) | 2 (40%) | 1.00 | **<0.05** |
| *R502C* | 1 | 0 (0%) | 1 (100%) | 1 (100%) | 0 (0%) | 1 (100%) | **< 0.001** | **< 0.001** |
| *T408M / R159W* | 1 | 1 (100%) | 0 (0%) | 1 (100%) | 1 (100%) | 0 (0%) | **< 0.001** | **< 0.001** |
| **Unknow** | 10 | 6 (60%) | 4 (40%) | 4 (40%) | 1 (25%) | 3 (75%) | **0.04** | **< 0.001** |
| *A495P* | 3 | 1 (33.3%) | 2 (66.6%) | 1 (33.3%) | 0 (0%) | 1 (100%) | **< 0.001** | **< 0.001** |
| *G154R / G232E* | 1 | 0 (0%) | 1 (100%) | 1 (100%) | 0 (0%) | 1 (100%) | **< 0.001** | **< 0.001** |
| *I528L* | 1 | 1 (100%) | 0 (0%) | 0 (0%) | 0 (0%) | 0 (0%) | **< 0.001** | - |
| *K13R* | 1 | 1 (100%) | 0 (0%) | 0 (0%) | 0 (0%) | 0 (0%) | **< 0.001** | - |
| *R78C* | 2 | 2 (100%) | 0 (0%) | 0 (0%) | 0 (0%) | 0 (0%) | **< 0.001** | - |
| *R83C* | 1 | 0 (0%) | 1 (100%) | 1 (100%) | 0 (0%) | 1 (100%) | **< 0.001** | **< 0.001** |
| *R83C / N409S* | 1 | 1 (100%) | 0 (0%) | 1 (100%) | 1 (100%) | 0 (0%) | **< 0.001** | **< 0.001** |
| **APOE ɛ4** | 23 | 8 (34.8%) | 15 (65.2%) | 12 (52.2%) | 3 (25%) | 9 (75%) | **0.002** | **< 0.001** |
| **APOE *ɛ4/ɛ4* (Hom)** | 4 | 1 (25%) | 3 (75%) | 2  (50%) | 0 (0%) | 2 (100%) | **< 0.001** | **< 0.001** |

^a^= comparison between GBA-PD F and GBA-PD M patients.

^b^= comparison between GBA-PD F (Cognitive Impairment) and GBA-PD M (Cognitive Impairment).

Significant p-values (p < 0.05) are reported in bold.
